# Hypertension Screening via Awake-Sleep Differences in Photoplethysmogram Signals

**DOI:** 10.1101/2025.05.22.25328194

**Authors:** Jingyuan Hong, Manasi Nandi, Yali Zheng, Jordi Alastruey

## Abstract

**Background:** Hypertension is a major risk factor for cardiovascular diseases. This study proposes a novel hypertension screening framework based on awake-sleep differences in photoplethysmography (PPG) indices, using machine learning. We hypothesised that normotensive individuals exhibit greater PPG variation between awake and sleep states than unmanaged hypertensive individuals.

**Methods:** The Aurora-BP dataset (n=180; 138 normotensive, 42 hypertensive) was used for model development, with 18 subjects reserved for internal testing. External validation was performed using the independent CUHK-BP dataset (n=26; 11 normotensive, 15 hypertensive). Twenty PPG-based indices were extracted, and subject-level p-values from Mann-Whitney U-tests comparing awake and sleep periods were used as model features. Discretised p-values served as inputs for four machine learning models.

**Results:** The Support Vector Machine (SVM) achieved the highest performance, with 81.1 ± 8.4% accuracy and 82.8 ± 8.1% F1-score on the internal test set using all indices. On the external test set, the SVM using only temporal indices achieved 84.6% accuracy and 86.7% F1-score. Temporal indices, especially those linked to the dicrotic notch, showed strong generalisability across datasets.

**Conclusion:** The study demonstrates the feasibility of awake-sleep PPG analysis for hypertension screening, highlighting the potential of wearable PPG devices for ambulatory monitoring.

**Novelty and Relevance:** *What Is New?:*

- This study introduces a novel machine learning–based hypertension screening method using awake-sleep differences in ambulatory photoplethysmogram (PPG) signals, validated on both internal and external datasets with a novel p-value–based feature selection strategy.

*What Is Relevant?:*

- The proposed approach enables non-invasive, cuff-free hypertension detection using wearable PPG data, offering improved generalisability and real-world applicability over traditional models.

*Clinical/Pathophysiological Implications?:*

- The ability to detect altered nocturnal cardiovascular patterns from PPG may support early identification of unmanaged hypertension and promote more accessible long-term monitoring.

## Introduction

Hypertension is a leading modifiable risk factor for cardiovascular mortality worldwide [1]. The standard method for diagnosing hypertension relies on absolute blood pressure (BP) values measured using an upper-arm cuff device, which operates via an inflatable cuff and an oscillometric algorithm to detect brachial BP readings [2]. Diagnosis strategies range from repeated office-visit readings to out-of-office monitoring and ambulatory measurements, depending on clinical guidelines and physician recommendations [2], [3]. Regular BP monitoring in daily life is also recommended for early detection and effective management of hypertension [2], [4]. However, current self-monitoring techniques, primarily cuff-based, can be uncomfortable and difficult to maintain over time, especially for frequent or ambulatory measurements [5], [6]. This highlights the need for a precise, user-friendly, and unobtrusive method for at-home hypertension screening and management.

Photoplethysmography (PPG), which measures peripheral blood volume changes during the cardiac cycle, is widely used in wearable devices [7]. PPG signals have been extensively studied as indicators of cardiovascular health, with applications ranging from atrial fibrillation detection to real-time BP estimation [8], [9]. Previous studies have used machine learning algorithms to explore the relationship between cardiovascular function and various PPG-based indices, including morphological, temporal, and derivative features [10], [11]. Despite its potential, PPG faces several challenges: the signal is sensitive to motion artefacts and contact pressure, and its stability can be affected by variations in amplitude and waveform shape across devices, populations, and environmental conditions [12], [13], [14]. As a result, many previous studies have been limited to single datasets, restricting the generalisability of their findings across diverse populations and settings [9], [15], [16], [17].

Ambulatory BP monitoring has been recognised in recent clinical guidelines as a more accurate method than office-based measurement. It provides comprehensive readings, reduces the risk of white coat hypertension, and minimally disrupts daily activities [2]. Crucially, it enables the detection of nocturnal BP patterns, which are key indicators of cardiovascular risk. Normotensive individuals typically exhibit a nocturnal ‘sleep-dip’ in BP, while hypertensive individuals may show altered patterns such as non-dipping, extreme dipping, or reverse dipping [18]. Nighttime readings are often more reliable due to reduced motion and temperature fluctuations [19]. Similarly, ambulatory PPG monitoring has the potential to capture awake-sleep variations in cardiovascular function [20], [21]. Continuous 24-hour PPG monitoring could detect these variations using a single wearable device, reducing the impact of measurement errors and inconsistencies across subjects, devices, and measurement conditions.

While ambulatory BP monitoring is well-established in current clinical guidelines [2], [3], [22], there remains a gap in methods that leverage less obtrusive physiological signals (i.e., PPG) for robust hypertension screening. Previous studies have focused on short-term or single PPG measurements within isolated datasets, limiting their clinical reliability and applicability. Moreover, fewer studies have examined whether diurnal variations in PPG indices can serve as indicators of cardiovascular health.

This study introduces a novel machine learning-based method for hypertension screening using ambulatory PPG measurements. We hypothesised that normotensive individuals would exhibit greater awake-sleep differences in PPG features, while individuals with unmanaged hypertension would show diminished or absent differences. To test this, we used two independent datasets: the Aurora-BP dataset (n=180) for model development and internal validation, and the CUHK-BP dataset (n=26) for external testing to assess generalisability.

## Methodology

### Dataset Description

The Aurora-BP dataset is an open-access resource containing data from 1,125 participants, collected to investigate cuff-based brachial BP readings, including systolic BP (SBP) and diastolic BP (DBP), along with Lead-I electrocardiogram (ECG), wrist PPG, and radial tonometric pressure waveforms [24]. Data were collected in two settings: laboratory (642 participants) and ambulatory (483 participants). PPG signals were recorded from the left wrist using an optical sensor (MAX30101, Analog Devices), while brachial BP was measured on the right arm using an ambulatory BP monitor (OnTrak 90227, Spacelabs Healthcare) (Figure 1a). During ambulatory monitoring, BP and PPG data were collected twice per hour during the daytime (8 a.m. to 8 p.m.) and once per hour at night (8 p.m. to 8 a.m.), producing 15-second PPG segments paired with single SBP and DBP measurements (Figures 1b and 1c) [24].

**Figure 1:**
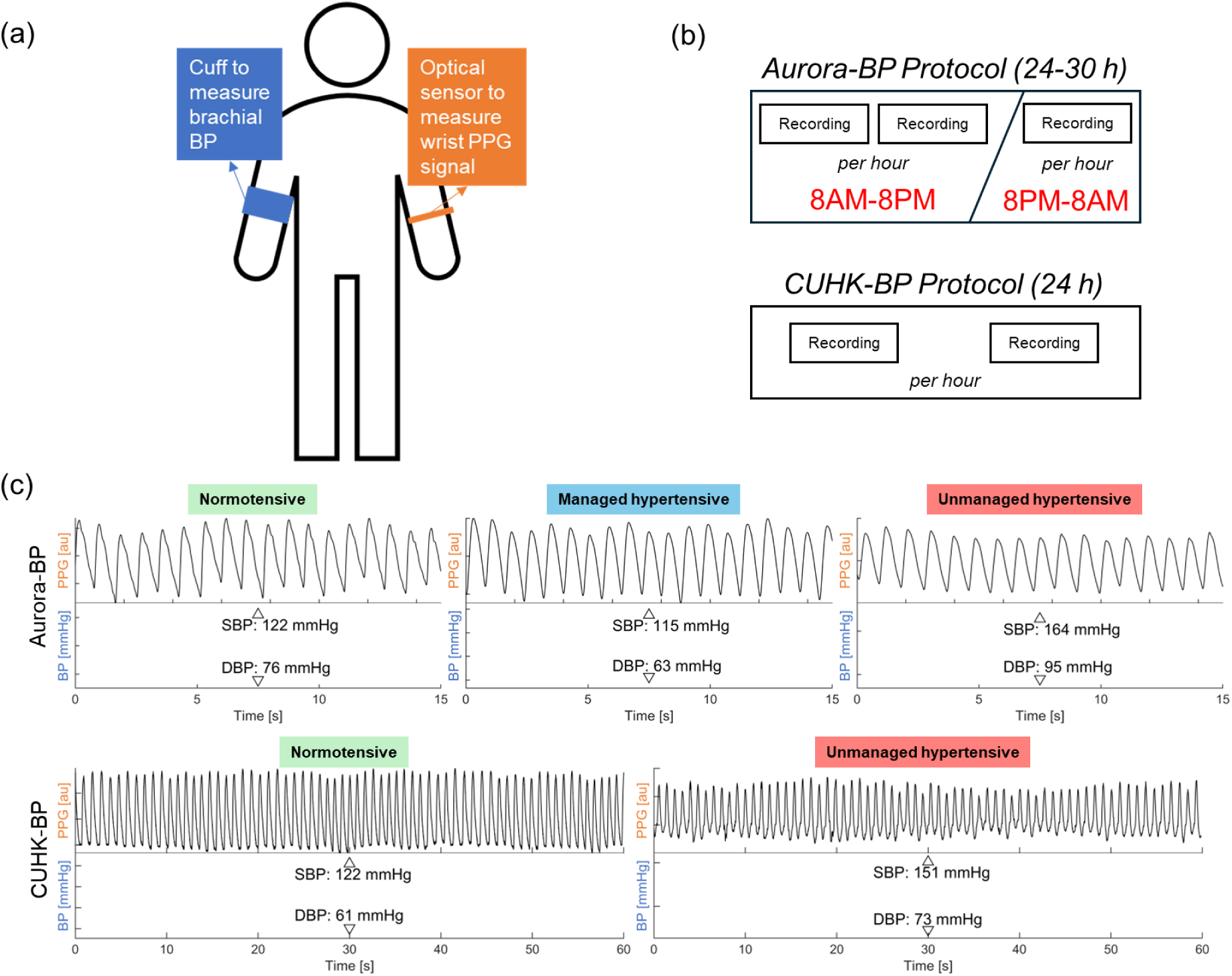
Ambulatory photoplethysmography (PPG) segments and blood pressure (BP) readings from the Aurora-BP and CUHK-BP datasets used in this study. (a) Data collection sites; (b) Measurement protocol; (c) Example PPG segments and corresponding BP readings from representative normotensive, managed hypertensive, and unmanaged hypertensive subjects in the Aurora-BP dataset (top) and CUHK-BP dataset (bottom).

From the ambulatory subset, 240 participants were included after preprocessing. This involved excluding segments with abnormal BP values (DBP < 30 mmHg) [25] and poor-quality PPG signals (quality score < 0.8) [24], reducing the sample from 483 to 441. Participants with fewer than three valid daytime and three valid nighttime measurements were further excluded, yielding a final cohort of 240. These participants were classified as normotensive, managed hypertensive, or unmanaged hypertensive (Table 1). Hypertensive status was self-reported based on the 2017 AHA guidelines [22], and classification labels were prelabelled in the dataset. Only the normotensive and unmanaged hypertensive participants were used for model training, testing, and evaluation as the internal dataset.

**Table 1:**
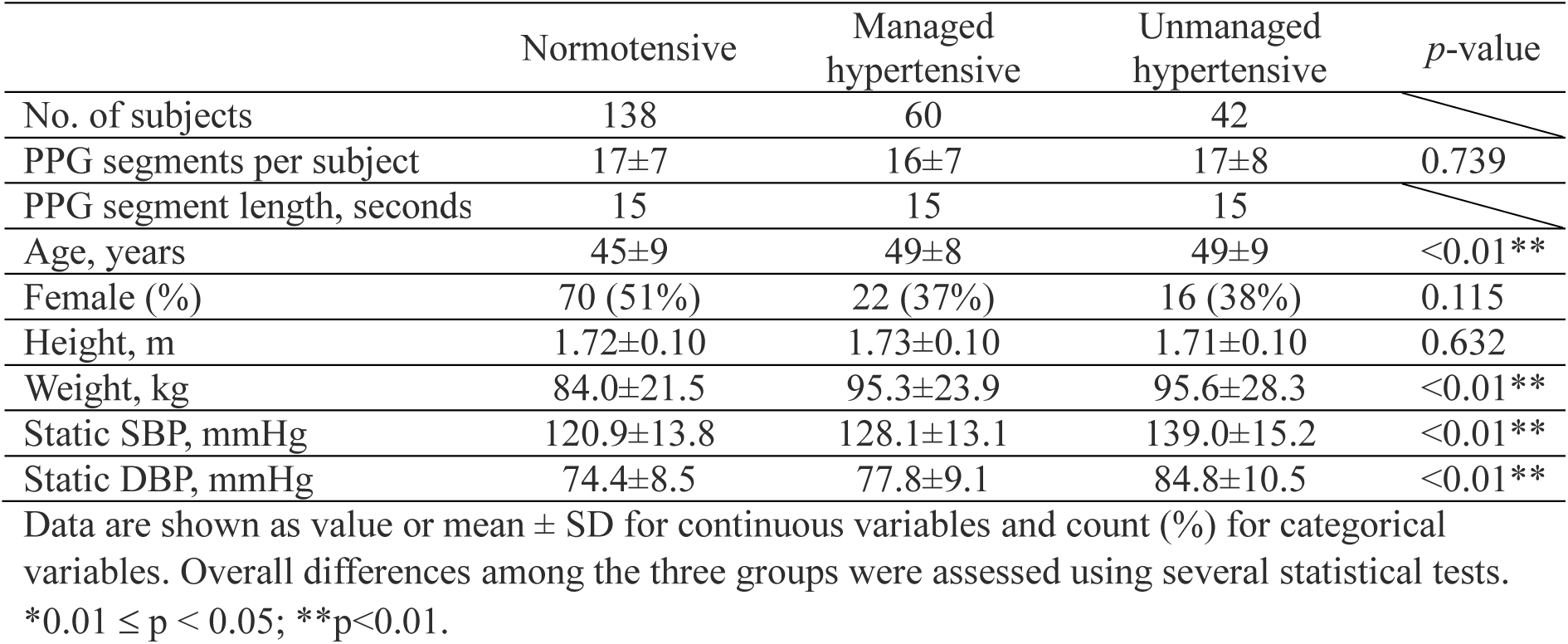
Characteristics of Aurora-BP cohorts used in this study.

The CUHK-BP dataset, collected by the Chinese University of Hong Kong and Prince of Wales Hospital [20], includes data from 26 participants who underwent 24-hour ambulatory monitoring using a commercial BP monitor (Oscar2, SunTech Medical) and a custom armband equipped with a near-infrared PPG sensor and ECG electrodes (two e-textile and one Ag-Ag/Cl). PPG signals were recorded from the left upper arm, and BP was measured on the right arm (Figure 1a). Data were collected twice per hour, yielding 60-second PPG segments with corresponding single SBP and DBP readings (Figure 1b and 1c). Demographic information is provided in Table 2. Participants were diagnosed as normotensive or unmanaged hypertensive according to the 2010 Hong Kong Reference Framework (HKRF) [26], with hospital-confirmed diagnoses. This dataset was used solely for external testing and evaluation of model generalisability.

**Table 2:**
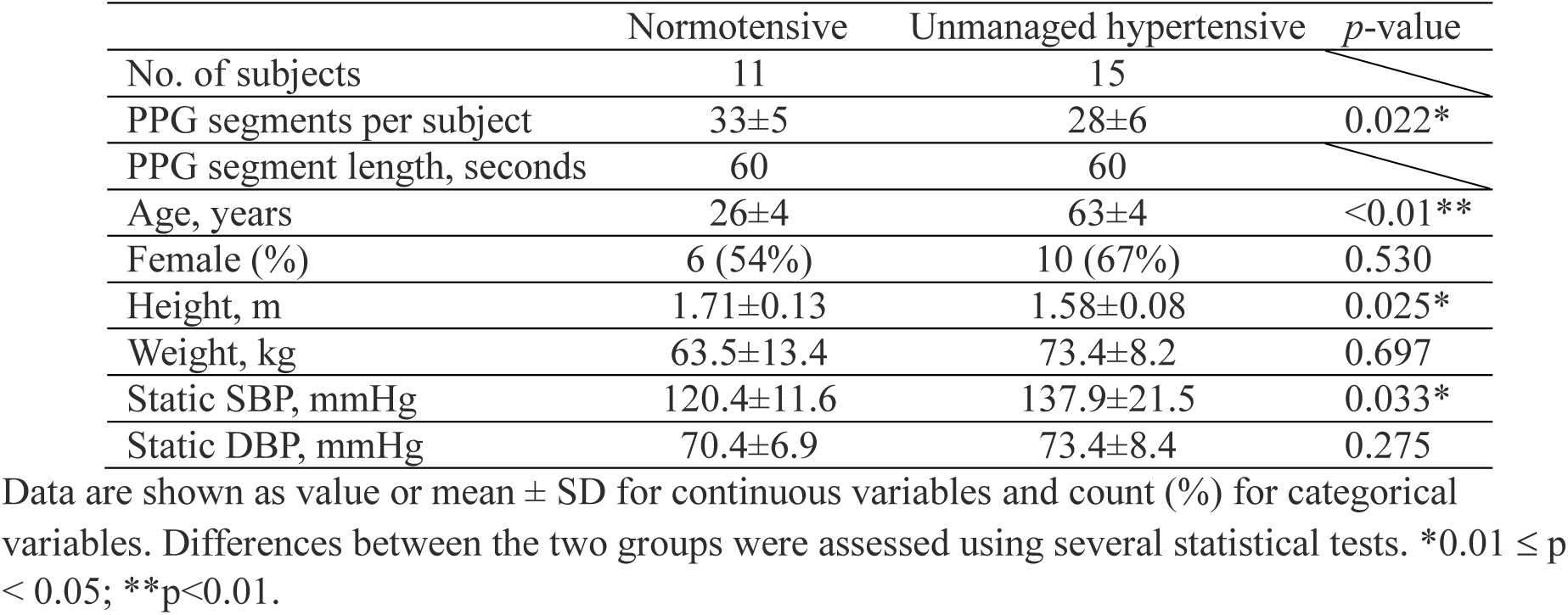
Characteristics of CUHK-BP cohorts used in this study.

### Signal Processing

The 60-second PPG segments in the CUHK-BP dataset were trimmed to 25 seconds by extracting the middle portion to remove unstable waveforms and noise typically found at the segment boundaries. Raw PPG segments from both datasets were then filtered using a 4th-order high-pass Butterworth filter (cut-off: 0.25 Hz) to eliminate low-frequency noise and baseline wander, followed by an equiripple low-pass finite impulse filter with 10 Hz passband, 12 Hz stopband, 1 dB passband ripple, and 60 dB stopband attenuation to suppress high-frequency noise (Figure 2a). Fiducial points were extracted from each beat within the filtered PPG segments using the PulseAnalyse algorithm [27] (Figure 2b). From these fiducials, 20 indices were computed as described in Supplementary Table S1. The average value of each index across all beats in a segment was then calculated for subsequent analysis.

**Figure 2:**
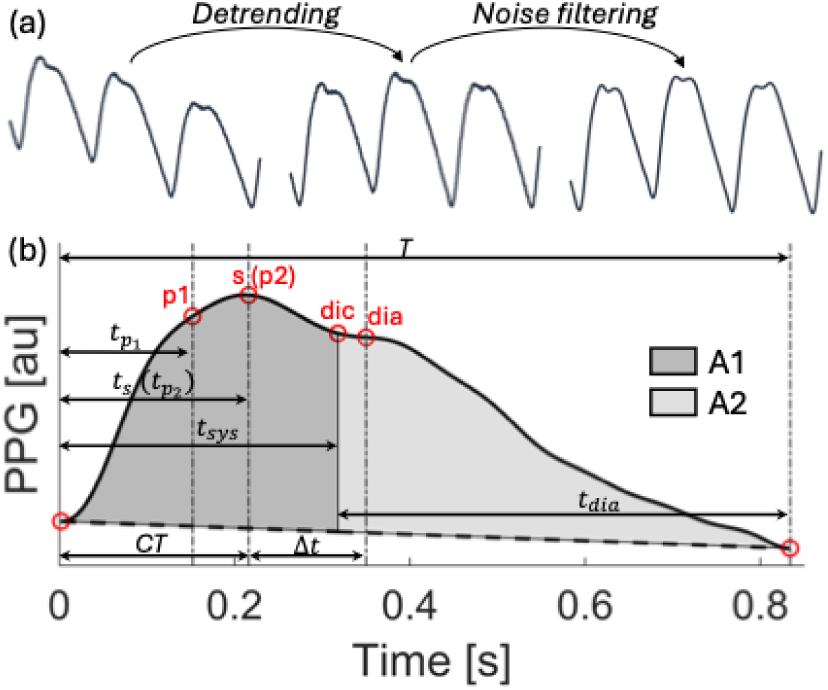
(a) Signal processing steps for PPG waveform pre-processing; (b) Illustration of the PPG fiducial points used to compute the PPG-based indices listed in Supplementary Table S1. All labels are defined in Table S1.

### Awake and Sleep Data Selection

The Aurora-BP dataset included step counts from the 30 minutes preceding each PPG segment recording. Sleep onset was defined as the first occurrence of zero steps after 8 p.m. (indicating sustained inactivity), and wake time as the first non-zero step count thereafter. These criteria delineated awake and sleep periods. In the CUHK-BP dataset, participant self-reported their sleep times, allowing direct separation of awake and sleep data.

### Statistical Analysis

Awake and sleep measurements were compared at both the cohort and individual levels.

At the cohort level, paired t-tests were used to assess awake-sleep differences in SBP, DBP, mean BP (MBP = (SBP+2*DBP)/3) [28], and pulse pressure (PP=SBP-DBP) across the three Aurora-BP cohorts and the two CUHK-BP cohorts. The difference between awake and sleep periods for all 20 PPG indices were also analysed using paired t-tests to assess statistical significance. Kruskal–Wallis tests were used to compare awake- and sleep-period SBP, DBP, MBP, PP, and continuous demographic variables among the three Aurora-BP cohorts, while Mann–Whitney U-tests were used for the same comparisons between the two CUHK-BP cohorts. Categorial variables were compared using chi-squared tests.

At the individual level, circadian SBP patterns were calculated as the percentage reduction from mean awake to mean sleep SBP and classified as dipper, extreme dipper, non-dipper, and reverse dippers [29]. Awake-sleep differences in the 20 PPG indices were evaluated for normotensive and unmanaged hypertensive subjects in both datasets. Due to the unequal numbers of awake and sleep measurements, Mann-Whitney U-tests were used. The resulting p-values reflected each index’s ability to distinguish between awake-sleep states while remaining robust to outlier values (particularly important given the noise sensitivity of PPG signals. To normalise and simplify machine learning model inputs, p-values were discretised into three categories: (i) highly statistically significant (p < 0.01) was set to 1, (ii) statistically significant (0.01 ≤ p < 0.05) was set to 0.5, and (iii) not statistically significance (p ≥ 0.05) was set to zero. These discretised set of 20 p-values were used as input features for machine learning models.

The frequency of significance labels was analysed across individuals within and between cohorts in the internal dataset to assess intra-cohort variability and inter-cohort separability.

### Machine Learning Models

Four binary classifiers—Logistic Regression (LR), K-Nearest Neighbours (KNN), Support Vector Machines (SVM), and eXtreme Gradient Boosting (XGBoost)—were used to classify subjects as normotensive or hypertensive. These models have previously been applied in hypertension screening studies [30], [31], [32].

The same processing pipeline, illustrated in Figure 3 was applied to each model. A randomly selected subset of 18 subjects (9 normotensive, 9 unmanaged hypertensive; 10% of Aurora-BP) was held out as an independent internal test set. The remaining 162 normotensive and unmanaged hypertensive subjects were used for training and validation. To address class imbalance (129 normotensive vs. 33 hypertensive), the Synthetic Minority Oversampling TEchnique (SMOTE) [33] was applied using five nearest neighbours to synthetically balance the dataset (157 normotensive, 159 hypertensive). The internal test set and the external CUHK-BP dataset were used for model evaluation.

**Figure 3:**
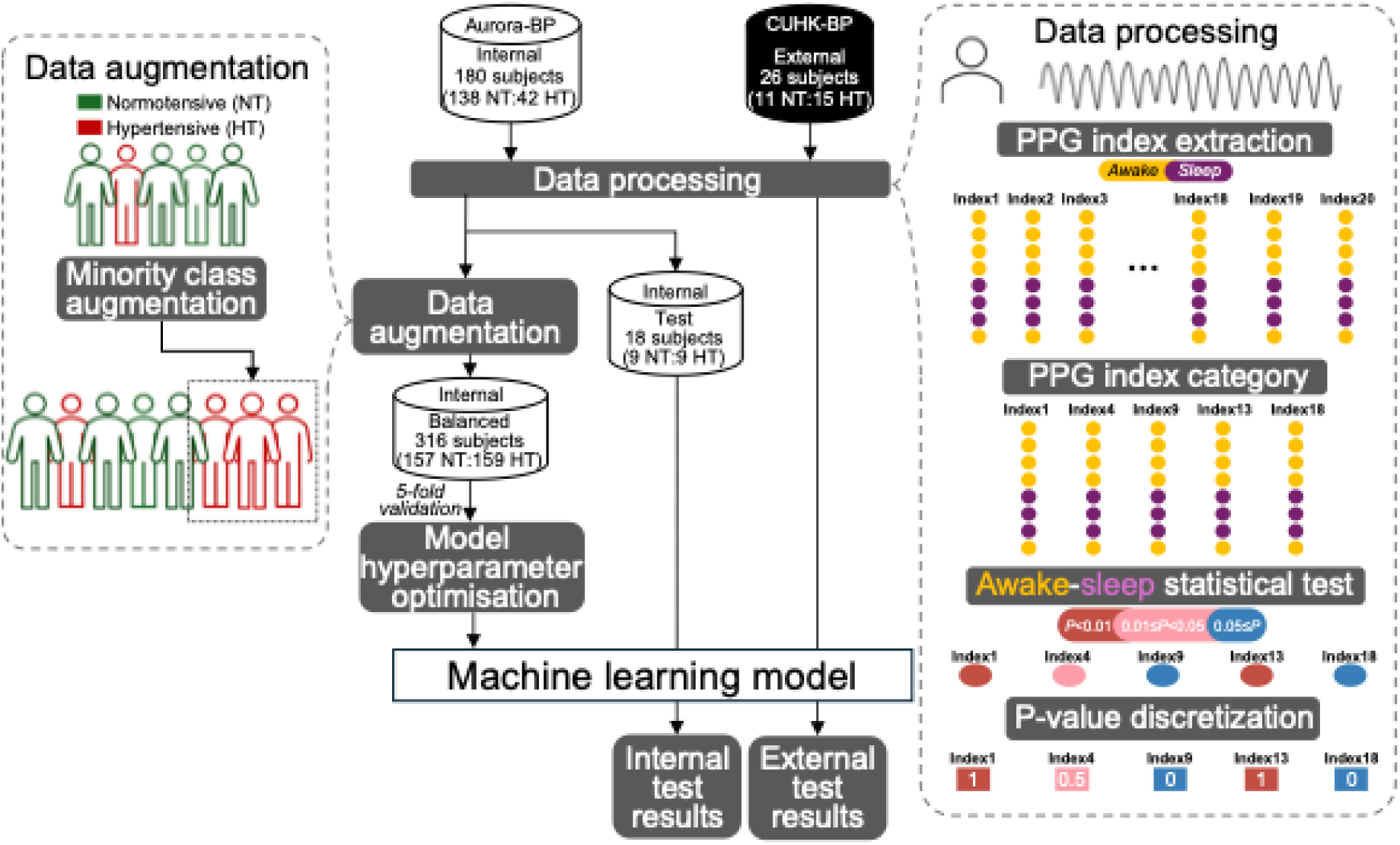
Flowchart of the model training and testing pipeline, including data preprocessing, feature extraction, class balancing, model training, hyperparameter optimisation, and evaluation on internal and external datasets.

All models were trained and optimised on discretised p-values from 20 PPG indices (11 temporal, 9 morphological) using the hyperparameters listed in Supplementary Table S2. A 5-fold cross-validation was used during training. Model performance was evaluated using accuracy and F1-score on both internal and external test sets. Internal testing included five random splits for robustness. For external testing, the model was trained on the entire internal dataset.

The 20 PPG-based indices were grouped into three categories: complete, temporal, and morphological. The complete group included all 20 indices listed in Table S1. The temporal group comprised 11 indices derived from time-fiducial points of the PPG waveform, while the morphological group included the remaining 9 indices extracted from morphological fiducial points.

The best-performing model was further analysed using SHapley Additive exPlanations (SHAP) [34] to evaluate the contribution of each index to classification decisions.

### Baseline Age-Only Model

Given that age is a well-known risk factor for hypertensive [35], a baseline model was developed for the best-performing binary classifier using age as the sole input feature. Due to the statistically significant age difference between normotensive and unmanaged hypertensive subjects in the CUHK- BP dataset (Table 2), this baseline model was trained and evaluated only on the internal Aurora-BP dataset to avoid confounding.

All signal processing, index extraction, and model development were implemented in MATLAB (version 2023a, MathWorks). A pseudocode representation of this process is provided in Figure S1.

## Results

### Awake and Sleep Differences

In the internal dataset, all three cohorts (normotensive, managed hypertensive, and unmanaged hypertensive) exhibited statistically significant reductions in cuff-based SBP, DBP, MBP and PP during sleep (Figure 4b). SBP, DBP and MBP followed similar circadian trends, reaching their lowest values around 2:30 a.m. (Figure 4a). Although PP changes were less visually pronounced (Figure 4a), they remained statistically significant (Figure 4b). The distribution of circadian SBP patterns (dipper, extreme-dipper, non-dipper, and reverse dipper) was similar across the three cohorts (Supplementary Table S3). However, comparisons across cohorts revealed significant differences in SBP, DBP, MBP and PP between the normotensive and unmanaged hypertensive groups during both awake and sleep periods (Supplementary Figure S3a).

**Figure 4:**
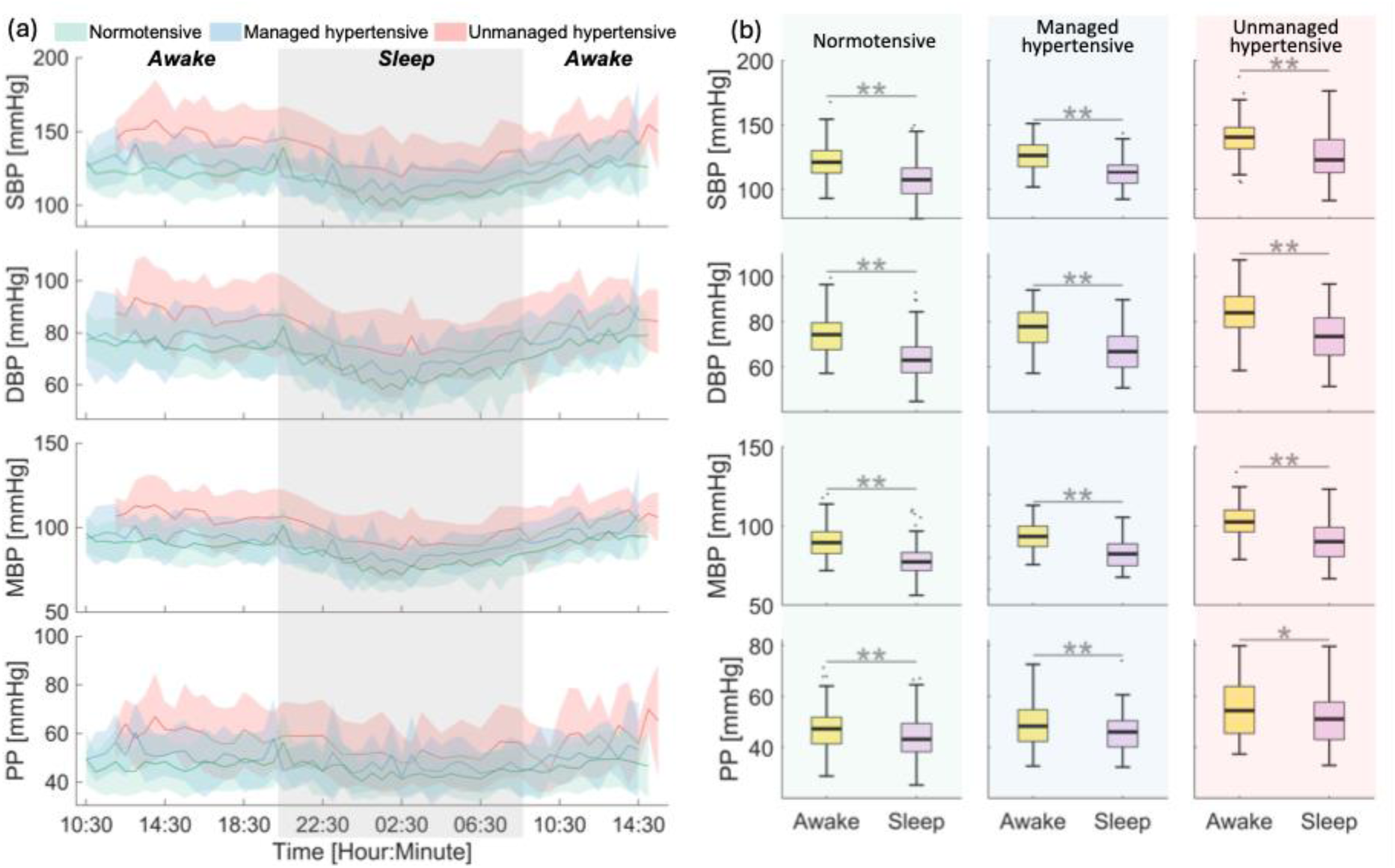
Blood pressure variations and statistical comparisons between awake and sleep periods across normotensive, managed hypertensive, and unmanaged hypertensive cohorts in the Aurora-BP dataset. (a) Trends in SBP, DBP, MBP, and PP across the full 24-hour ambulatory measurement period. Solid lines represent cohort means and shaded areas indicate standard deviations. (b) Statistical comparisons of SBP, DBP, MBP and PP between awake and sleep periods for each cohort.

In the external dataset, DBP and MBP also decreased significantly during sleep, while SBP showed a significant reduction only in the unmanaged hypertensive cohort (Supplementary Figure S3). PP showed no significant change. These findings align with SBP patterns distributions (Table S3), where the normotensive cohort showed a low proportion of dippers and reverse dippers (9.1% each) and a high proportion of non-dippers (81.8%), whereas the unmanaged hypertensive cohort had more dippers (33.3%) and reverse dippers (20.0%). All measures except for SBP and PP during sleep showed significant differences between normotensive and unmanaged hypertensive groups (Supplementary Figure S3b).

PPG waveform comparisons in Figure 5 further illustrate typical awake-sleep differences. In normotensive individuals, the awake-period waveform exhibited a clear dicrotic notch and diastolic peak, which were less prominent during sleep and in hypertensive subjects. Multiple PPG-derived indices also showed significant awake-sleep differences, particularly in the normotensive group (Table 3). Three temporal indices (𝑡_𝑠𝑦𝑠_, 𝑡_𝑑𝑖𝑎_, 𝑡_𝑝1−𝑑𝑖𝑎_) and three morphological indices (*RI*, 𝑟𝑎𝑑𝑖𝑜_𝑝2−𝑝1_, and *A2*) were highly significant in normotensives (p < 0.01), but not in hypertensives (p ≥ 0.05). Four additional temporal indices (Δ𝑡, SI, 𝑝𝑟𝑜𝑝_𝑠_, 𝑡_𝑟𝑎𝑑𝑖𝑜_) and two morphological indices (𝑅𝐼_𝑝2_ and 𝑟𝑎𝑡𝑖𝑜_𝑝2−𝑝1_) show moderate significance in normotensives (0.01 ≤ p < 0.05), but again were not significant in hypertensives. The managed hypertensive cohort showed a p-value distribution more closely aligned with the normotensive group, differing in only five indices.

**Figure 5:**
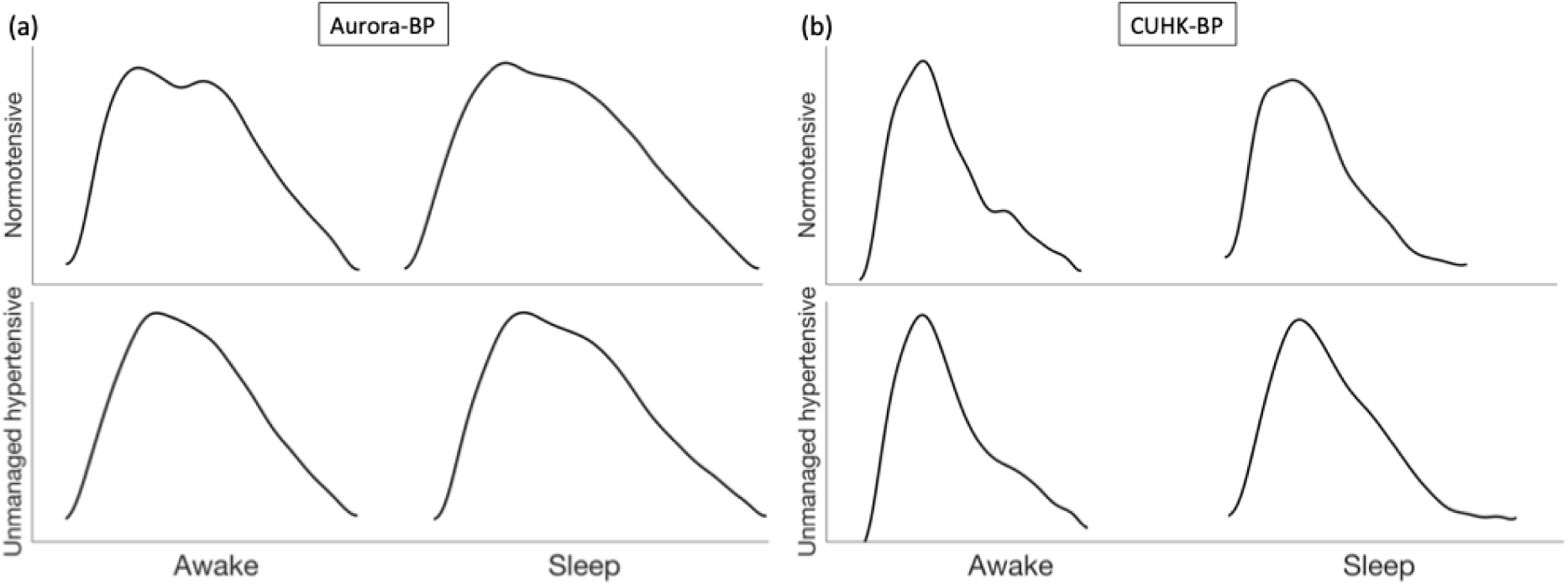
Example PPG waveforms recorded during awake and sleep periods in normotensive (top) and unmanaged hypertensive (bottom) individuals from the (a) Aurora-BP and (b) CUHK-BP datasets.

**Table 3:** Statistical comparison of awake and sleep PPG indices across the three Aurora-BP cohorts.

| Index | Normotensive | Managed hypertensive | Unmanaged hypertensive |
| --- | --- | --- | --- |
| $\Delta t$ | * | - | - |
| $CT$ | ** | ** | ** |
| $SI$ | * | - | - |
| $CT/h$ | ** | ** | ** |
| $prop_s$ | * | * | - |
| $t_{sys}$ | ** | ** | - |
| $t_{dia}$ | ** | ** | - |
| $t_{ratio}$ | * | * | - |
| $prop_{\Delta t}$ | ** | ** | * |
| $t_{p_1-dia}$ | ** | ** | - |
| $t_{p_2-dia}$ | - | - | - |
| $AI$ | ** | ** | ** |
| $AP$ | ** | ** | ** |
| $RI$ | ** | ** | - |
| $RI_{p_1}$ | ** | * | * |
| $RI_{p_2}$ | * | * | - |
| $ratio_{p_2-p_1}$ | ** | - | - |
| $A1$ | ** | ** | * |
| $A2$ | ** | ** | - |
| $IPA$ | - | * | - |

A dash (-) indicates no statistical significance; \* denotes statistical significance ( $0.01 \leq p < 0.05$ ); \*\* indicates high statistical significance ( $p < 0.01$ ).

Although most indices were not statistically significant across all subjects (Figure 6), normotensive individuals showed a higher proportion of significant indices than unmanaged hypertensives (8.3% vs. 3.2%). Notably, several indices (𝐶𝑇/ℎ, 𝑡_𝑠𝑦𝑠_, 𝑡_𝑑𝑖𝑎_, 𝑡_𝑝1−𝑑𝑖𝑎_, 𝐴𝐼, 𝐴𝑃, 𝑅𝐼_𝑝1_, 𝑟𝑎𝑡𝑖𝑜_𝑝2−𝑝1_ and 𝐴1) were highly significant (p < 0.01) in over 10% of normotensive subjects.

**Figure 6:**
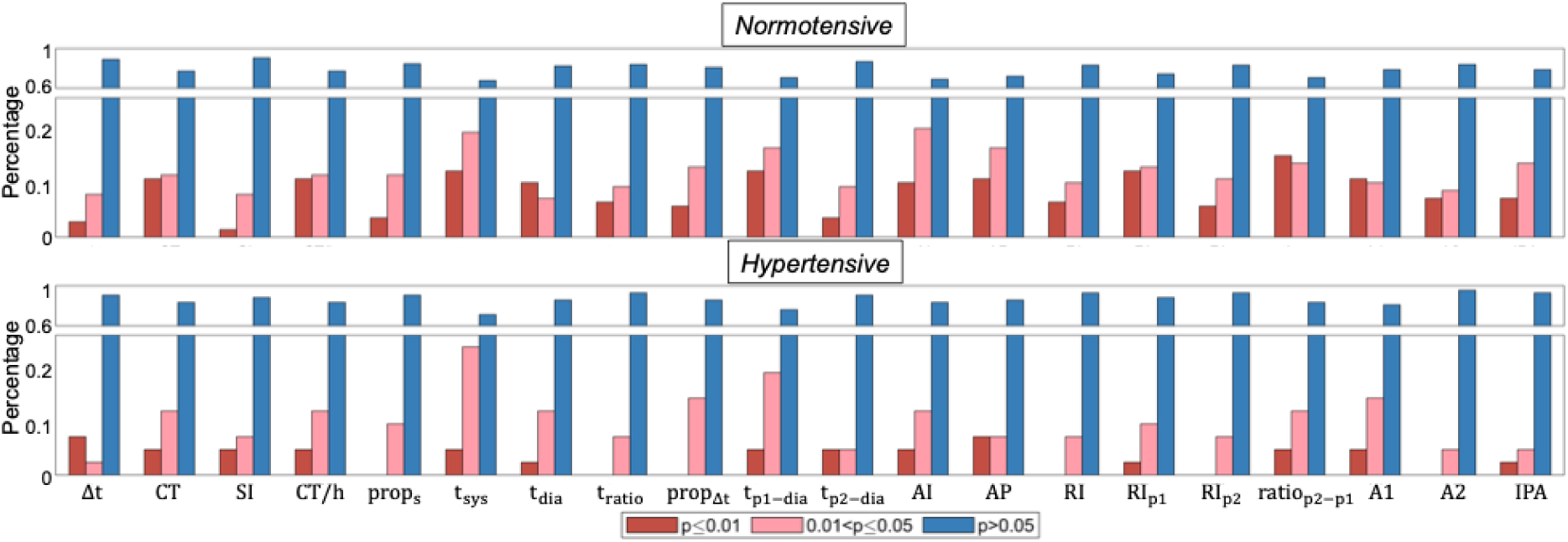
Frequency distribution of p-value ranges for all 20 PPG-based indices in normotensive (top) and unmanaged hypertensive (bottom) individuals from the internal Aurora-BP dataset. Highly significant values (p < 0.0) are shown in red, significant values (0.01 ≤ p < 0.05) in pink, and non-significant values (p ≥ 0.05) in blue.

### Hypertension Screening Performance

The SVM model achieved the best performance on both datasets, although with different sets of indices (Table 4). On the internal dataset, it attained the highest accuracy (81.1% ± 8.4%) and F1- score (82.8% ± 8.1%) using all 20 indices. On the external dataset, it reached an accuracy of 84.6% and an F1-score of 86.7% using only temporal indices. LR and XGBoost also performed well externally with temporal indices, each achieving accuracy and F1-scores above 70% and 80%, respectively. In contrast, KNN consistently showed the weakest performance, with internal scores below 63% and external scores below 45%. Overall, the complete index set was most effective internally, while temporal indices yielded better generalisability externally.

**Table 4:** Classification performance of the four models on internal and external test datasets using different categories of PPG-based indices.

|  | Index category | Logistic Regression |  | K-Nearest Neighbours |  | Support Vector Machines |  | eXtreme Gradient Boosting |  |
| --- | --- | --- | --- | --- | --- | --- | --- | --- | --- |
|  |  | Accuracy | F1-score | Accuracy | F1-score | Accuracy | F1-score | Accuracy | F1-score |
| Internal dataset | Complete | 65.6%<br>(4.7%) | 71.1%<br>(2.0%) | 63.3%<br>(5.0%) | 61.2%<br>(4.2%) | **81.1%**<br>**(8.4%)** | **82.8%**<br>**(8.1%)** | 81.1%<br>(6.3%) | 82.1%<br>(6.2%) |
|  | Temporal | 65.6%<br>(6.1%) | 69.0%<br>(6.8%) | 53.3%<br>(3.0%) | 29.6%<br>(5.5%) | 72.2%<br>(8.8%) | 73.8%<br>(9.8%) | 71.1%<br>(4.7%) | 71.2%<br>(7.1%) |
|  | Morphological | 62.2%<br>(8.2%) | 70.0%<br>(5.4 %) | 48.9%<br>(8.2%) | 23.3%<br>(11.4%) | 65.6%<br>(8.2%) | 69.2%<br>(7.5%) | 72.2%<br>(11.8%) | 77.6%<br>(8.0%) |
| External dataset | Complete | 69.2% | 69.2% | 42.3% | 44.4% | 57.7% | 52.2% | 61.5% | 61.5% |
|  | Temporal | 73.1% | 75.9% | 50.0% | 38.1% | **84.6%** | **86.7%** | 80.8% | 82.8% |
|  | Morphological | 73.1% | 74.1% | 38.5% | 27.3% | 61.5% | 58.3% | 61.5% | 58.3% |

In the best-performing SVM model, internal testing showed a higher true negative rate (0.91) than true positive rate (0.71), with a false negative rate of 0.29 and a false positive rate of 0.09. Performance on the external dataset was more balanced across all classification metrics (Figure 7).

**Figure 7:**
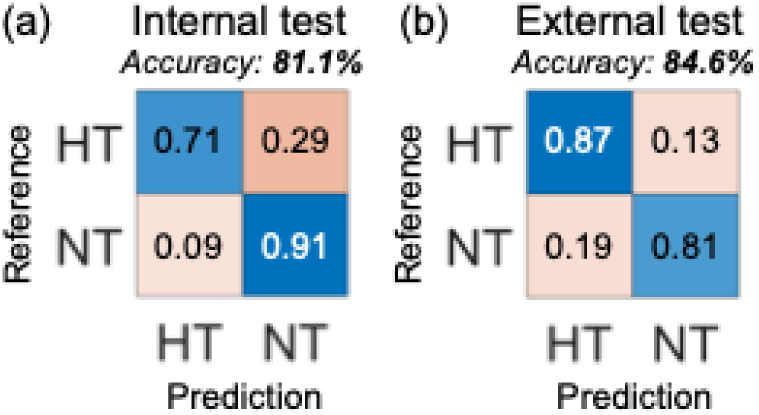
Best classification results of the Support Vector Machines model on (a) the internal Aurora-BP test dataset and (b) the external CUHK-BP test dataset. HT: hypertensive; NT: normotensive.

SHAP analysis identified four morphological and six temporal indices among the top 10 most important features for hypertension classification in the internal test using all 20 indices (Figure 8a). In the external test, the most important indices were 𝑡_𝑑𝑖𝑎_, 𝑡_𝑝1−𝑑𝑖𝑎_, 𝑡_𝑝2−𝑑𝑖𝑎_, 𝑡_𝑠𝑦𝑠_, and 𝑡_𝑟𝑎𝑡𝑖𝑜_; all related to the dicrotic notch (Figure 8b).

**Figure 8:**
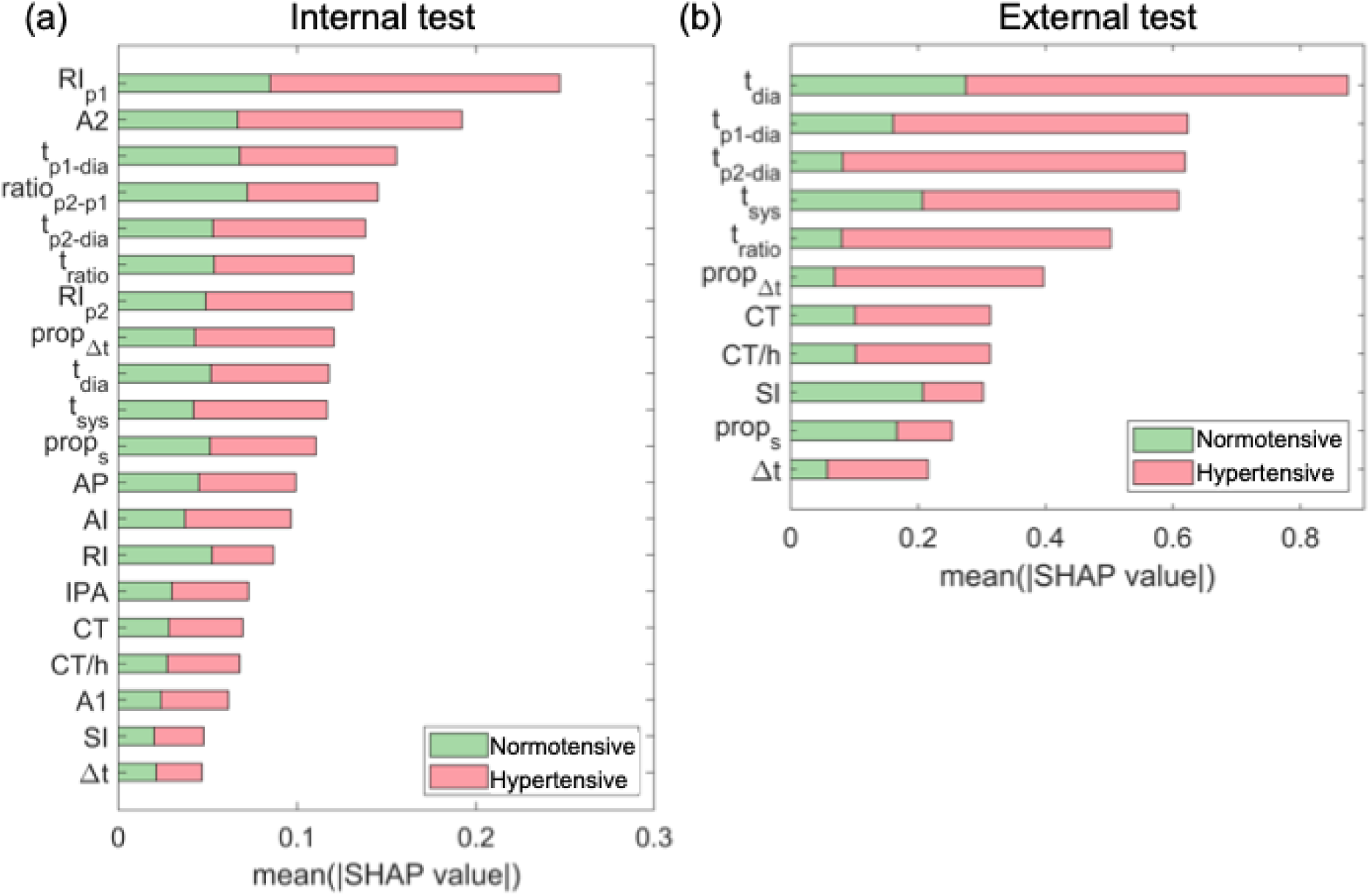
SHapley Additive exPlanations (SHAP) values computed using the Support Vector Machines model for (a) the complete set of PPG-based indices on the internal (Aurora-BP) test dataset and (b) the temporal indices on the external (CUHK-BP) test dataset. Green bars represent the mean absolute SHAP values contributing to normotensive predictions; red bars represent those contributing to hypertensive predictions.

The baseline SVM model using age alone achieved 67.8% ± 12.7% accuracy and a 70.5% ± 10.7% F1-score on the internal dataset, indicating that age was a moderate predictor but less effective than PPG-based indices.

Compared to previous studies, our p-value-based SVM approach achieved competitive accuracy and F1-scores on both internal and external datasets (Table 5), outperforming methods in [36] and [30], and approaching those in [31], [36] and [37], which lacked external validation.

**Table 5:** Comparison of the proposed method with previous studies. Each study is characterised by the input type (1st column), training dataset and sample size (2nd column), test dataset and sample size (3rd column), machine learning model used (4th column), accuracy (5th column), and F1-score (6th column).

| Input | Training | Test | Model | Accuracy | F1-score |
| --- | --- | --- | --- | --- | --- |
| 17 PPG indices [39] | 16 subjects (private dataset) | 1 subject (private dataset) | AdaBoost |  | 63% |
| 10 PPG indices [31] | 80 MIMIC subjects | 80 MIMIC subjects | KNN |  | 86.9% |
| 4 PPG indices [30] | 140 MIMIC III recordings | 140 MIMIC III recordings | SVM | 72.9% | 77.0% |
| A PPG index [36] | 30 subjects (private dataset) | 13 subjects (private dataset) | Multiple instance learning | 85.5% | 89.2% |
| 24 PPG indices [37] | 405 Aurora-BP subjects | 405 Aurora-BP subjects | Causality graphs + LR | 89.0% | 87.0% |
| 20 complete PPG indices (this study) | 162 Aurora-BP subjects | 18 Aurora-BP subjects | Circadian statistical analysis + SVM | 81.1% (8.4%) | 82.8% (8.1%) |
| 11 temporal PPG indices (this study) | 180 Aurora-BP subjects | 26 CUHK-BP subjects | Circadian statistical analysis + SVM | 84.6% | 86.7% |

## Discussion

This study demonstrated the feasibility of hypertension screening using ambulatory PPG measurements through a novel machine learning framework that leverages awake-sleep differences in PPG morphology. Analysis of two independent datasets revealed distinct circadian variations in PPG-derived indices between normotensive and unmanaged hypertensive individuals. The support vector machine model achieved the highest classification performance, with approximately 85% accuracy and F1-score on an external dataset unseen during training. Temporal indices proved particularly robust across datasets, highlighting their generalisability. Our novel p-value-based approach matched or outperformed previous methods and, notably, included external validation; highlighting its potential for non-invasive, continuous hypertension screening in ambulatory settings.

Previous studies have shown that normotensive individuals typically exhibit a nocturnal BP dip, while hypertensive individuals may display altered patterns [18]. Our internal data confirmed this dip in normotensive participants but showed similar circadian trends, with no substantially altered patterns, in both hypertensive groups. Despite comparable BP patterns across cohorts, 13 of the 20 PPG indices exhibited highly significant awake-sleep differences in the normotensive cohort, compared to only four in the unmanaged hypertensive group. This suggests that PPG captures distinct physiological responses to sleep not reflected by BP alone. The managed hypertensive cohort showed PPG patterns more closely aligned with the normotensive group, indicating that treatment may partially restore normal cardiovascular function [39]. Notably, although BP-based sleep-awake differences were less pronounced in the external dataset, PPG-based indices still effectively distinguished normotensive from unmanaged hypertensive individuals, underscoring the sensitivity of our novel p-value-based approach to subtle sleep-related cardiovascular changes.

At the individual level, both normotensive and hypertensive groups exhibited many non-significant awake-sleep differences. However, normotensives had a higher frequency of highly significant changes, reflecting greater physiological variability and supporting the use of machine learning to leverage these differences for classification.

Temporal indices yielded the strongest external performance, suggesting they capture hypertension-related features that are more generalisable than morphological indices, which may be more dataset-specific. These findings are consistent with previous work demonstrating the robustness of temporal features across PPG sensors and anatomical sites [11].

Unlike other studies that included demographic variables such as age [40], we excluded them to avoid confounding, as normotensive and hypertensive cohorts in the external dataset differed significantly in age. This allowed for a focused assessment of PPG-derived features. While age was a strong predictor of hypertension in our internal dataset, our PPG-based model substantially outperformed an age-only baseline.

Among the tested models, SVM consistently outperformed others across datasets. KNN, while moderately effective internally, performed poorly externally, likely due to its reliance on distance-based similarity metrics, which are sensitive to data distribution shifts. In contrast, SVM, XGBoost and LR, which learn underlying patterns, maintained stronger cross-dataset performance. SVM’s advantage may stem from its focus on critical samples near the decision boundary rather than fitting the entire dataset to find optimal decision boundaries [41], making it more robust in small, imbalanced datasets. The internal dataset’s imbalance led SVM to favour normotensive predictions during internal testing, while SVM applied to the external dataset using temporal indices produced more balanced classification results.

SHAP analysis identified the most influential features for classification. In the internal dataset, 𝑅𝐼_𝑝1_ and *A2* (both morphological indices) ranked highest, consistent with their known association with arterial stiffness and vascular ageing; factors implicated in disrupted diurnal BP variation in hypertension [42]. These differences in diurnal index variation reflect underlying vascular pathophysiology. Temporal indices were more influential in the external dataset. A previous ICU-based study using 24-hour measurements found that the circadian rhythm of 𝑡_𝑠𝑦𝑠_, 𝑡_𝑟𝑎𝑡𝑖𝑜_, and *CT* correlated more with SBP than heart rate, while Δ𝑡 showed no such correlation [43]. Our findings aligned with these results, except for *CT* which showed moderate importance across both groups. Notably, the top five ranked indices (𝑡_𝑑𝑖𝑎_, 𝑡_𝑝1−𝑑𝑖𝑎_, 𝑡_𝑝2−𝑑𝑖𝑎_, 𝑡_𝑠𝑦𝑠_, and 𝑡_𝑟𝑎𝑡𝑖𝑜_) are all related to the dicrotic notch of the PPG waveform, which marks aortic valve closure in the cardiac cycle. Sleep typically lowers heart rate and delays valve closure in normotensives, a pattern often disrupted in hypertension [44], [45], [46]. These disruptions may explain the higher importance of dicrotic-notch-related indices in our SHAP analysis and their strong generalisability.

This study has several limitations. First, both datasets were relatively small. Despite this, our method achieved performance comparable to or better than previous work using larger datasets, many of which lacked external validation. Second, we focused on binary classification, whereas clinical guidelines define multiple hypertension stages [2], [3], [22]. Third, we excluded managed hypertensive subjects from classification due to their absence in the external dataset. Future research should consider multiple hypertension stages and expand to larger, more diverse ambulatory datasets to further enhance the generalisability, reliability and clinical relevance of this approach.

## Conclusion

This study introduced a novel, machine learning-based hypertension screening method that leverages awake-sleep differences in PPG indices. This approach effectively distinguished normotensive from hypertensive individuals and demonstrated strong generalisability on an independent dataset. Temporal indices, particularly those related to the dicrotic notch, proved more robust across datasets than morphological indices. By using a p-value-based approach, our method overcomes common challenges such as PPG signal instability and noise sensitivity, thereby expanding the clinical utility of PPG. To our knowledge, this is the first study to exploit awake–sleep PPG variations for hypertension assessment, highlighting the potential of wearable devices for non-invasive, ambulatory monitoring in real-world settings.

## Data Availability

The Aurora-BP dataset is publicly available. All models, training code, and data preprocessing scripts will be made publicly available to support reproducibility and further development.

